# Modelling the potential impact of mask use in schools and society on COVID-19 control in the UK

**DOI:** 10.1101/2020.09.28.20202937

**Authors:** J. Panovska-Griffiths, C.C. Kerr, W. Waites, R.M. Stuart, D. Mistry, D. Foster, D.J. Klein, R.M. Viner, C. Bonell

**Affiliations:** Department of Applied Health Research, University College London, London, UK; Institute for Global Health, University College London, London, UK; The Queen’s College, Oxford University, Oxford, UK; Institute for Disease Modeling, Seattle, WA, USA; School of Physics, University of Sydney, Sydney, NSW, Australia; School of Informatics, University of Edinburgh, UK; Department of Mathematical Sciences, University of Copenhagen, Copenhagen, Denmark; Disease Elimination Program, Burnet Institute, Melbourne, VIC, Australia; Rethink Priorities, Redwood City, CA, USA; UCL Great Ormond St. Institute of Child Health, London, UK; Faculty of Public Health and Policy, London School of Hygiene and Tropical Medicine, London, UK

**Keywords:** COVID-19, masks, test-trace-isolate strategies, mathematical modelling, agent-based model

## Abstract

Recent findings suggest that an adequate test-trace-isolate (TTI) strategy is needed to prevent a secondary COVID-19 wave with the reopening of society in the UK. Here we assess the potential importance of mandatory masks in the parts of community and in secondary schools. We show that, assuming current TTI levels, adoption of masks in secondary schools in addition to community settings can reduce the size of a second wave, but will not prevent it; more testing of symptomatic people, tracing and isolating of their contacts is also needed. To avoid a second wave, with masks mandatory in secondary schools and in certain community settings, under current tracing levels, 68% or 46% of those with symptomatic infection would need to be tested if masks’ effective coverage were 15% or 30% respectively, compared to 76% and 57% if masks are mandated in community settings but not secondary schools.

## Introduction

Evidence to date suggests that SARS-CoV-2, the coronavirus that causes COVID-19, is mainly transmitted when someone who has COVID-19 coughs, sneezes or exhales and releases droplets of infected fluid [1]. Some of the droplets can be breathed in by people within a close proximity and some will fall on nearby surfaces and objects. If people touch the contaminated objects and then touch their eyes, nose or mouth, COVID-19 may also be transmitted [2-3]. Reducing the frequency of physical contact, maintaining good hygiene and good ventilation, and pursuing effective test-trace-isolate (TTI) strategies are important non-pharmaceutical interventions that can reduce COVID-19 transmission before a vaccine or effective anti-viral drugs become available.

In the early stages of the COVID-19 pandemic, there was uncertainty over the effectiveness of face coverings in reducing the spread of COVID-19 and protecting the public [4-5]. While public use of face coverings was adopted early in many Asian countries that have had experience with epidemics such as SARS in 2003 [6], countries such as the USA and the UK were slower in mandating face coverings, despite relatively high levels of public support [7]. For the purposes of this study, we will refer to face coverings or masks interchangeably to mean face protection that covers the mouth and nose.

There is now considerable evidence supporting the effectiveness of masks for protecting against transmission between individuals. Laboratory experiments have found that almost all types of masks can greatly reduce droplet emission and viral shedding by infectious wearers [8-9] suggesting their effectiveness for source control. Two observational studies [10-11] and recent systematic reviews focusing on SARS-1, MERS and influenza [12-15] indicate that masks also substantially reduce infection risk to the non-infected wearer, even when their infectious contact is unmasked. Specifically, Chu et al. [12] suggest that face coverings use could result in a reduction in infection risk of around 44% (95% CI = 11–60%) in a community setting, with stronger associations in a healthcare setting and with use of N95 respirators compared to surgical or cotton masks. No randomised controlled trials (RCTs) have been completed on face coverings against COVID-19 or other coronaviruses (although one is currently underway [16]). Observational studies are prone to confounding, and RCTs focused on influenza are inconclusive [13]. Epidemiological studies have also shown a negative association between mask prevalence and COVID-19 incidence at a city, state and national level [17-20].

The use of face coverings is becoming mandatory in certain community settings across the four UK nations. In England, masks became mandatory on public transport on June 15, 2020, and in shops from July 24, 2020, with further expansion of places where masks are mandatory announced on July 31, 2020 [21] and new rules being enforceable by law from August 08, 2020. In Scotland, masks have been mandatory on public transport since June 22, 2020 and in shops since July 10, 2020 [22]. Masks have been mandatory on public transport in Wales and Northern Ireland since July 10, 2020 [23].

However, the magnitude of the effect of compulsory masking on COVID-19 incidence is still highly uncertain since there is no straightforward way to infer a change in transmission probability from a change in viral emission. Mathematical modelling, having already played an important role in informing policy around the COVID-19 pandemic [24-31], can help to assess the likely impact of compulsory masking. Models has been used to evaluate the impact of the lockdown [29-30], to explore the efficacy of different test-trace-isolate (TTI) strategies [26-28,31] and a growing number of modelling studies considering the impact of masks on the COVID-19 epidemic [32-39]. While the overall message from these models is that, as lockdown measures are relaxed, masks are likely to be effective if they are worn by a large percentage of the population, they often optimistically assumed that a large proportion of transmission events could be prevented with face masks by overestimating their efficacy and guided by evidence of the impact of masks on influenza transmission reduction [15]; hence their results are likely to overestimate impact. Furthermore, with the exception of a few studies [36-39], many studies to date have used population-based models that do not differentiate between household, school, workplace and community contacts. Under the current policy of mandatory masks wearing in the UK, only the community contacts will be affected and untangling different layers of contacts is crucial in evaluating the impact of masks. In summary, existing modelling studies are likely to exaggerate the impact of face coverings.

Our recent work [28], which used the detailed individual-based model called Covasim [36], highlighted that adequate TTI is needed to prevent a secondary epidemic wave when broader society including schools reopen in the UK. Here, we extend this work by calibrating Covasim, illustrated in Figures 1A-B, to recent data, to August 28, 2020 rather than June 17, 2020 as in [28], taking in consideration the slower-than-anticipated reopening of society in the UK during July and August, and to explore whether extending mandatory mask use to secondary school students alongside its existing use in some community settings could contribute to reducing the risk of COVID-19 wave resurgence later this year. The most recent findings from the CoMIX studies suggest that the contact rate in England is increasing much slower than anticipated with an average of 4 contacts per person during July and August compared to around 11 contacts per day in pre-COVID-19 era [30]. We anticipate that this will increase with reopening of schools but less than the anticipated 90% of the pre-COVID-19 contact rate previously modelled in [28].

**Figure 1:**
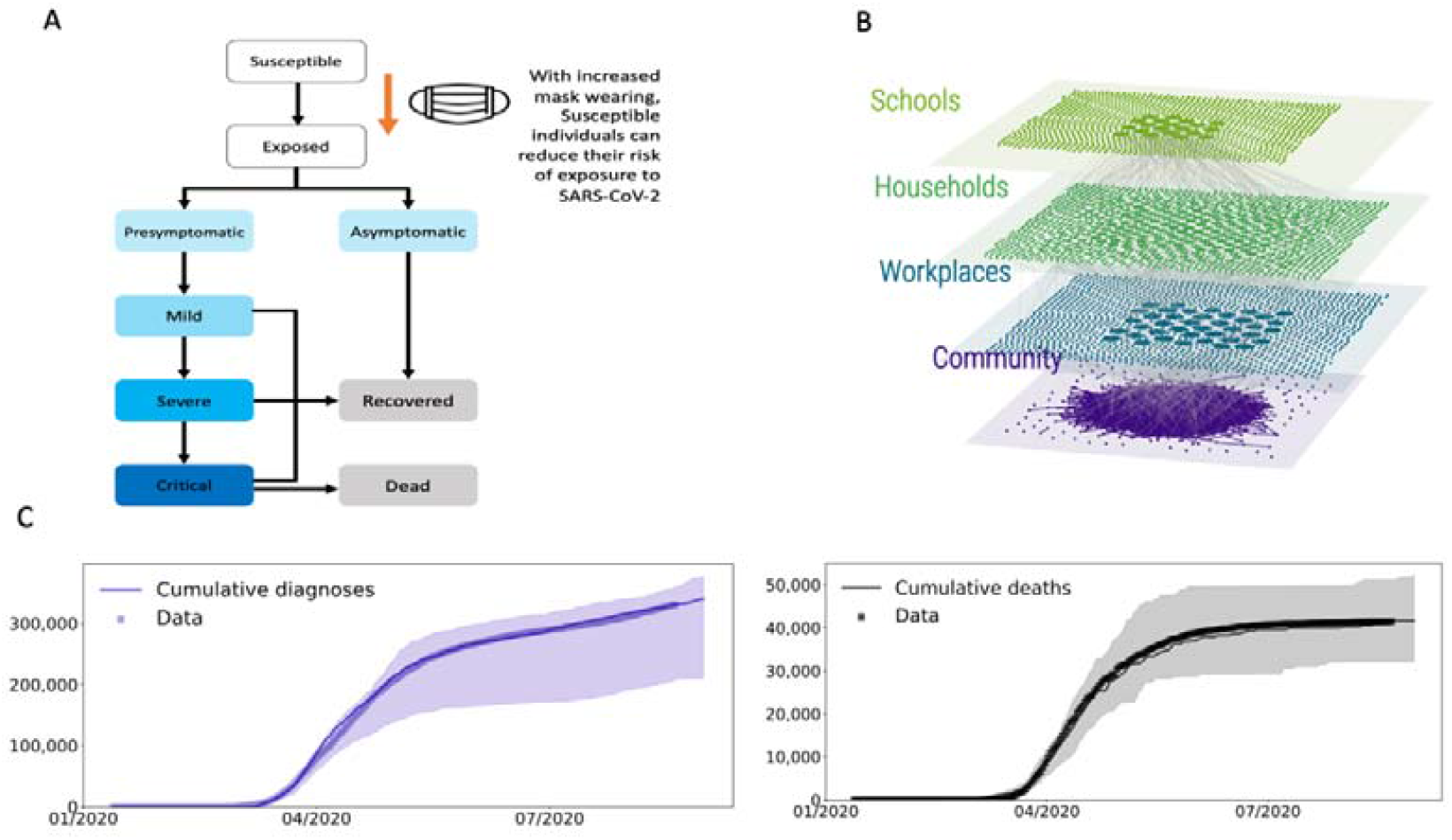
A: Schematic of the model component showing the modelled effect from using face coverings. B: Illustration of the different levels modelled in Covasim C: Results of the model calibration showing the matching of the model projected cumulative COVID-19 cases and cumulative deaths associated with COVID-19 with the data from https://coronavirus.data.gov.uk. Data is shown in thick blue/black lines, medians across twelve simulations are indicated by thin blue/blue lines and 10% and 90% quantiles by blue/grey shading.

WHO recommends mask use for children over 12 years under the same conditions as adults, with decisions on mandatory use for those between 6-11 based on a set of conditions [40]. This recommendation is on the basis that younger children may have lower susceptibility and potentially lower transmissibility than adults [41]. While the evidence is not conclusive, we anticipate the practicalities with younger children correctly obeying masks rules are more challenging than for older children. In addition, face masks are more likely to hamper the education of primary than secondary school children given the focus on learning basic speaking and social skills. Children over 12 years old in the context of the UK school systems attend secondary schools and recently, following WHO guidance [42], the government has permitted schools to mandate mask wearing in communal areas (where high levels of social mixing and lower levels of social distancing may occur) [43]. Overall data on mask use in children is sparse, with a recent WHO document [40] suggesting that mask fit and compliance is likely to be poorer in children than in adults and hence mask efficacy levels based upon adult data may need to be adjusted. To account for this uncertainty in the compliance with mask-wearing and their efficacy, we simulate two levels of masks’ effective coverage in schools and community settings (estimated as the product of the masks efficacy (per-contact risk reduction) and coverage (the proportion of contacts in which they are worn); see Methods). We aim to estimate the different combinations of testing and tracing levels necessary to avoid COVID-19 resurgence after September 2020 and a second COVID-19 wave later this or next year, accounting for community mask-wearing and scenarios with and without mandatory masks in secondary schools, including two levels of masks’ effective coverage. The strategies we have explored have been discussed with members of scientific advisory bodies in the UK.

## Results

As part of calibrating the model, we estimated the daily probabilities of testing people with symptoms between January 21, 2020 and August 28, 2020. These were 2.77% for both July and August, corresponding to around 24% of people with symptomatic infection tested during their illness (assuming an average symptomatic period of around 10 days). We note that this is higher than the 18% testing level when we calibrated until June 17, 2020 in [28]. Furthermore, in calibrating to the UK epidemic, we estimated that 1,500 people were infected in the UK on January 21, 2020, that the per-contact transmission probability was 0.72% under the assumption that 70% of infections were symptomatic, and that 0.75% of those with asymptomatic COVID-19 infections were tested at some point during their illness (see Methods for details). Figure 1C shows the results of the model calibration.

The projections from the calibrated model across all scenarios are shown in Figures 2-4. In Figure 2 we show the heatmaps of the peak of the cumulative infections for different trace (x-axis) and test (y-axis) levels. Increases in cumulative infections result from resurgences of COVID-19; comparable increases can be seen in new infections and deaths (Figures S1-S2). In Figure 2, higher cumulative infections are shown in darker shades of red, while lower values are lighter colours. Across these figures (and respectively Figures S1-S2 of the supplementary material), a very light orange colour represents a region within the heatmaps where the resurgence of COVID-19 is controlled with combinations of adequate test-trace and mask wear.

**Figure 2:**
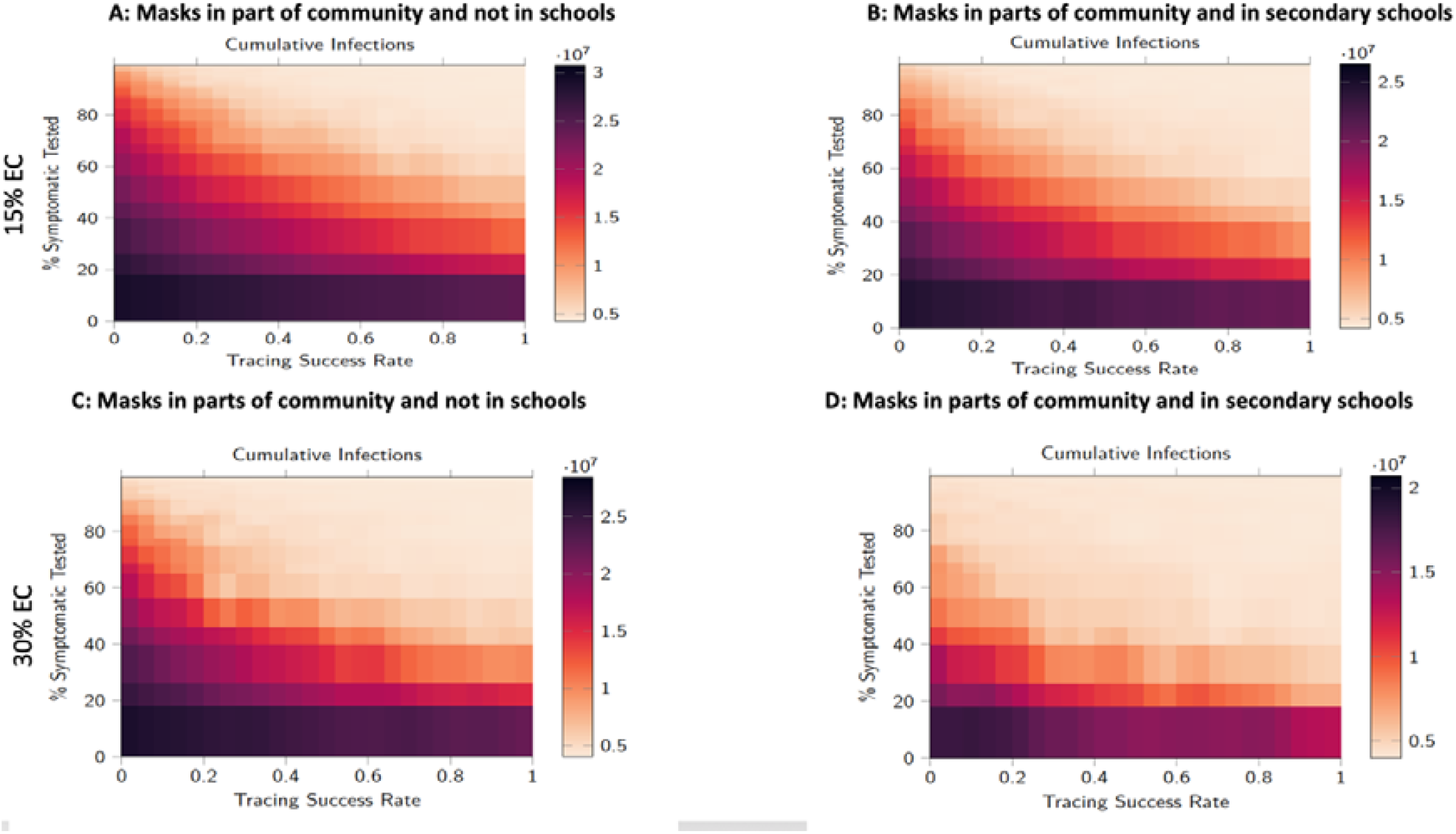
Heatmaps of cumulative infections for different trace (x-axis) and test (y-axis) levels across the scenario of masks wear in parts of community with and without schools masks’ wear. Higher cumulative infections are shown in darker shades of red, while lower values are lighter colours with a light orange colour representing a region within where the resurgence of COVID-19 is controlled with combinations of adequate test-trace and mask usage.

**Figure 3:**
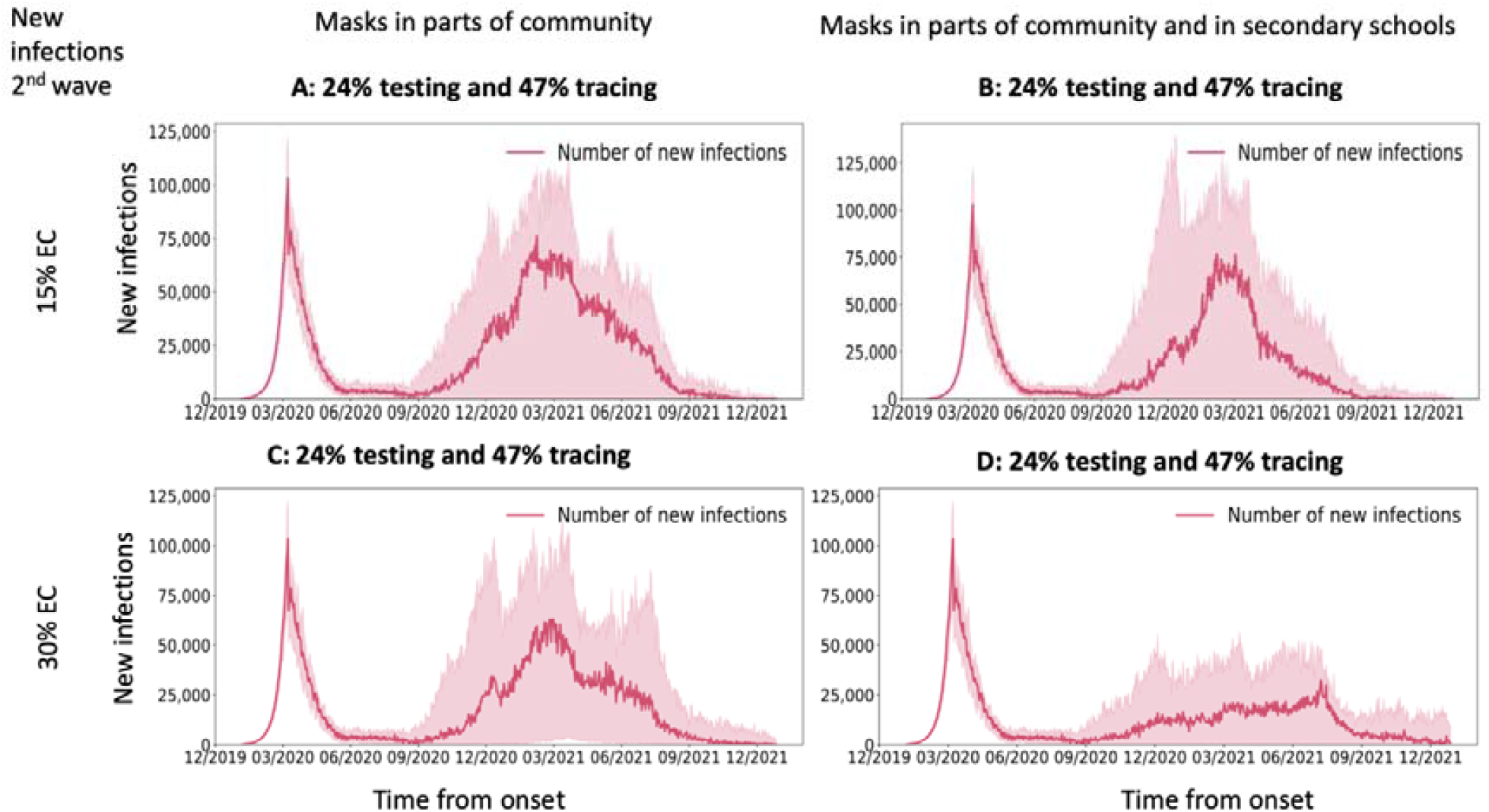
Model scenarios of potential second COVID-19 epidemic wave in the UK during 2020-2021 under different policies of masks’ wear and their effective coverage for testing level of 24% and tracing level of 47%. Medians across twelve simulations are indicated by solid red lines and 10% and 90% quantiles by red shading. The results do not change if we run a larger number of simulations and we tested 10, 12, 20 and 30 simulations. The difference is that the noise in the simulations increases with increased sample of simulations and this is why we chose twelve simulations for the figures here.

**Figure 4:**
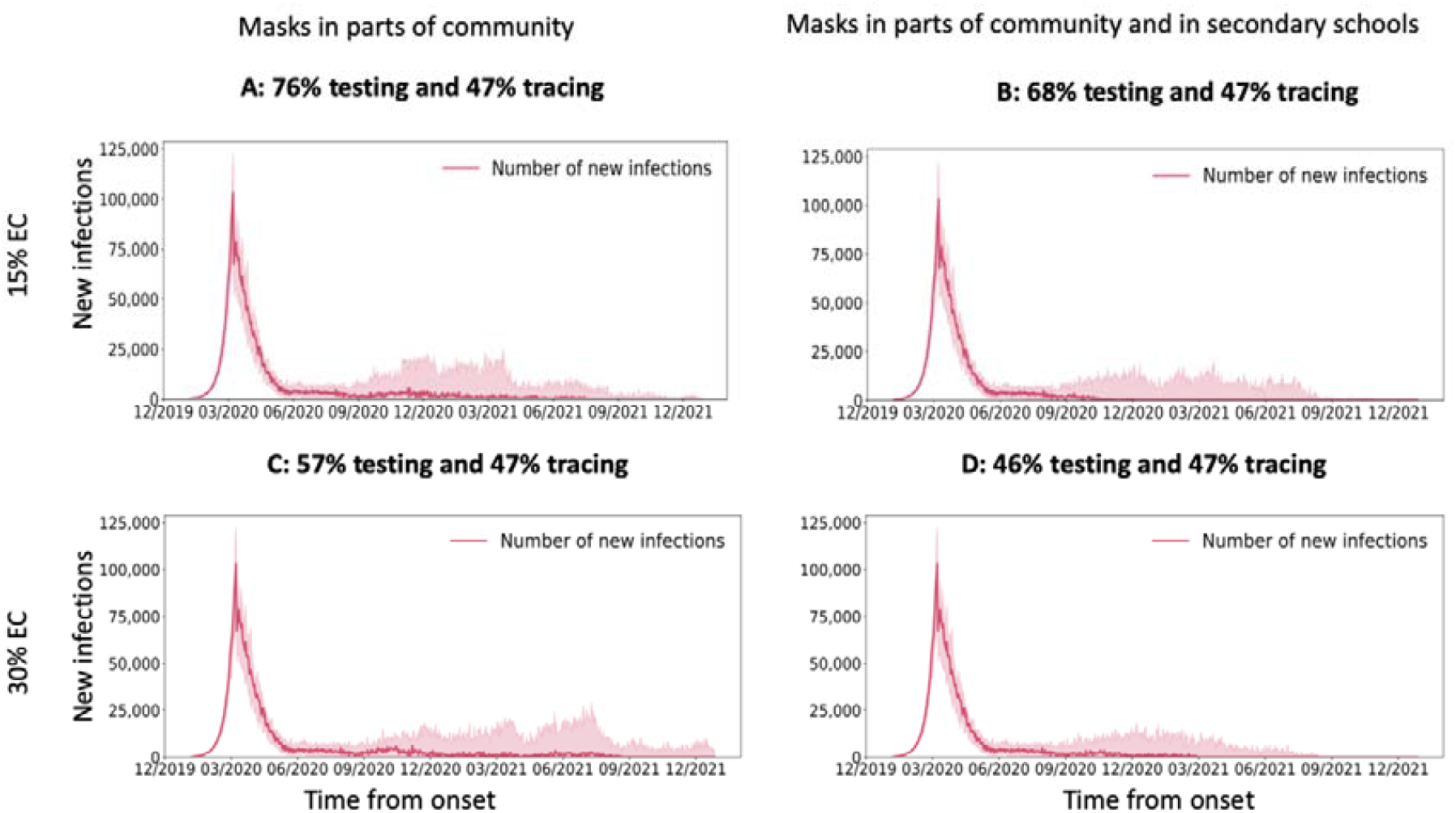
Model scenarios of potential second COVID-19 epidemic wave in the UK during 2020-2021 under different policies of masks’ wear and their effective coverage for testing and tracing levels where future the resurgence of COVID-19 is prevented. Medians across twelve simulations are indicated by solid red lines and 10% and 90% quantiles by red shading. The results do not change if we run a larger number of simulations and we tested 10, 12, 20 and 30 simulations. The difference is that the noise in the simulations increases with increased sample of simulations and this is why we chose twelve simulations for the figures here.

In Figures 3-4, across the four scenarios, we show illustrative temporal profiles for the new infections for two different combinations of test-trace levels: the current 24% testing and 47% tracing level with predicted second wave and a test-trace combination where the resurgence of COVID-19 is avoided.

### Masks worn in some community settings but not in secondary schools

Reopening of broad areas of society in the UK together with schools from September 01, 2020, with masks mandatory in parts of community from July 24, 2020 but not in schools, will result in an increase in COVID-19 cases if test-trace levels are insufficient under both assumptions about masks’ effective coverage (dark red region in Figures 2A and 2C). Under the current testing level (24% of symptomatic people tested at some point of their infection) and with 47% of contacts traceable, we predict an increase in the number of new COVID-19 infections from September 2020, which in these scenarios peak around March 2021, under both assumptions of masks’ effective coverage (Figure 3A and 3C). The strength of the secondary COVID-19 wave is predicted to vary depending on the effective coverage of masks (Figure 3A and 3C), but could be up to the same level as the original COVID-19 wave in the UK.

With adequate combinations of test-trace levels, a second epidemic wave may be avoided (Figure 2A and 2C, light orange-coloured region). For example, to achieve this, assuming 47% of contacts could be traced, as was the case in July and August 2020, with low mask effective coverage (15%) in relevant community settings, 76% of those with symptomatic infection would need to be diagnosed and isolated (Figure 4A). If masks’ effective coverage in community settings were higher (30%), the necessary testing level would be 57% (Figure 4C).

### Masks worn in secondary schools alongside some community settings

With reopening of broad areas of society in the UK together with schools from September 01, 2020, with masks mandatory in parts of community from July 24, 2020 but also in secondary schools, our model predicts a secondary COVID-19 wave will again occur unless there is an adequate test-trace program. This is the case under both assumptions of masks’ effective coverage (dark red region in Figures 2B and 2D). For the scenario of low (15%) effective coverage of masks, under current testing (24%) and tracing levels (47%), the model predicts that a resurgence in COVID-19 cases is likely to start from September 2020 and peak around March 2021 (Figure 3B). However, with higher (30%) effective coverage of masks, and if they are mandatory in secondary schools, the strength of the second epidemic wave is predicted to be much less. This is evident from comparing the size of the light orange region in Figures 2B and 2D and also when comparing Figures 3B and 3D for current test-trace levels.

With a scaled-up version of the current TTI strategy, the resurgence in COVID-19 can be avoided (Figures 4B and 4D). To achieve this, assuming current tracing levels (47% of contacts traced and isolated) continue in future, testing levels of 68% or 46% of those with symptomatic infection during their infectiousness period are necessary if masks’ effective coverage is respectively 15% or 30%.

### Masks in secondary schools vs masks not in secondary schools

Our results suggest that there is a greater benefit of mandatory masks in secondary schools if the effective coverage of masks is high (30%) (Figure 3A vs 3B and 3C vs 3D). Under current testing and tracing levels (24% testing, 47% tracing) and masks’ effective coverage of 30%, the predicted second COVID-19 wave would be less than half of the original wave if masks were mandatory in secondary schools as well as used in community settings (comparing Figures 3C and 3D). If the effective coverage of masks is less (15%), the effect of the mask wearing in schools on the predicted wave is much less (comparing Figures 3A and 3B). The minimum testing levels necessary to avoid a second wave, under scaled up TTI, is 8-11% less when masks are mandatory in schools than if they are not, depending on the effective coverage of masks (76% and 57% in Figures 3A and 3C compared to 68% and 46% in Figures 3B and 3D respectively).

## Discussion

### Summary of findings

Our results suggest that, with broader society including schools reopened from September 2020, and with current levels of coverage of TTI, mandating the use of masks in secondary schools would result in fewer infections, but would not be sufficient to prevent a secondary COVID-19 wave in the UK later this year. Only with increased TTI coverage could a secondary COVID-19 wave be avoided. The necessary TTI coverage requires testing sufficient number of symptomatic people and then sufficient identification, tracing and isolation of their positive contacts. Across different assumptions of masks’ effective coverage, different levels of testing and tracing would be necessary to avoid a second epidemic wave. For example, if masks were mandatory in secondary schools, assuming that current tracing levels of 47% continue, 68% or 46% of those with symptomatic infection would need to be tested respectively under scenarios of 15% and 30% mask effective coverage. If masks were not mandatory at secondary schools, the respective numbers would be 76% and 57% for 15% and 30% effective coverage of masks in the relevant community settings.

### Novelty of our work, policy relevance and putting in context of current literature

Overall, our findings suggest that making masks mandatory in secondary schools would be of some benefit, but that scaling up of TTI coverage is necessary to prevent resurgence of COVID-19 later this year and beyond while we await an effective vaccine. We highlight that adoption of masks in schools, in addition to using in community settings, can help reduce a potential secondary epidemic wave, but, to do this effectively, the effective coverage of masks has to be assumed to be high (30% in our case). If it were lower, the reduction in the potential second wave would be smaller. Uncertainties concerning the effectiveness of the masks remain, and results add to the ongoing body of evidence on the impact of using face masks against the epidemic spread.

Unlike previous work that has considered specific values of testing level and projecting outcomes, our modelling provides all combinations of thresholds for testing and tracing coverage that could prevent a COVID-19 resurgence and a secondary wave whether or not mask were mandatory in secondary schools. Furthermore, previous studies [32-39] have explored the broader impact of masks by simulating lowered transmission across all layers in society. The Covasim model instead has the granularity to consider specific layers, hence allowing our study to specifically explore the impact of mandatory masks in secondary schools, in combination with different levels of TTI coverage.

### Limitations of our work

The analyses presented here have several limitations. First, while we have made an effort to model the UK epidemic, some of the parameters we have used are from other settings. Secondly, as with any modelling study, we have made a series of assumptions within the modelling framework, for example concerning the proportion of infections which are symptomatic, and the susceptibility and infectiousness of children compared to adults. Large uncertainty regarding the proportion of asymptomatic infection remains with recent evidence suggesting that asymptomatic incidence has a wide range of 2-57%. In our model, we have assumed that symptomatic infections account for 70% of onward-transmitted infections, as a number of other models have assumed [26-29] and that development of symptoms is age-dependent analogous to other studies [26-29].

The model considers mask wearing to be uniform in the sense that, for a given level of coverage in a subpopulation (say a school), the average transmission probability is reduced by the same amount. In reality, there are four possible cases for an interaction between a susceptible and an infectious individual: both are wearing a mask, neither are wearing a mask, or one or the other is wearing a mask. The transmission probability will be different for each of these four cases. Because the way in which transmission probability can be expected to vary is not well-understood, we use ‘effective coverage’ as a catch-all for the four different cases. More research is required to understand the extent to which these fine-grained details matter.

We also note that we have set the level of contact tracing to be the same across all contact network layers. We are aware that this may lead to oversimplification and that some contact network layers may be more likely to be traced than others. For example, the reports from NHS Test and Trace [50] summarised in Table 1 until end of August, report % contacts reached by complex (e.g. schools and workplaces) and non-complex cases (e.g. households) suggesting that complex cases have better tracing as they are usually in institutions where contacts identities are better known. The percentage of contacts reached within the community are not reported and we expect that challenges associated with finding, tracing and isolating are with contacts in the community, and that this may reduce the overall impact of the programme. With this in mind, within the model we have assumed an average contact tracing level across all layers.

**Table 1:** Levels of contact tracing during June 2020, July 2020 and August 2020 collated from weekly report from NHS Test and Trace available from [50].

| *Weeks* | *% of people tested positive that are reached within 1-2 days* | *% of people tested positive that provided 1 or more close contacts* | *% of close contacts reached within 1-2 days* |  |
| --- | --- | --- | --- | --- |
| *28<sup>th</sup> May – 24<sup>th</sup> June* | 73.9 | 67.6 | 86.4 | [50] |
| *25<sup>th</sup> June -1<sup>st</sup> July* | 77.4 | 75.8 | 70.8 | [50] |
| *2<sup>nd</sup> July – 8<sup>th</sup> July* | 78.7 | 78.2 | 71.1 | [50] |
| *9<sup>th</sup> July – 15<sup>th</sup> July* | 79.7 | 79.9 | 77.9 | [50] |
| *16<sup>th</sup> July – 22<sup>nd</sup> July* | 81.4 | 81.3 | 75.1 | [50] |
| *23<sup>rd</sup> July -29<sup>th</sup> July* | 79.4 | 79.7 | 72.4 | [50] |
| *30<sup>th</sup> July – 5<sup>th</sup> August* | 79.7 | 78.6 | 71.4 | [50] |
| *6<sup>th</sup> August -12<sup>th</sup> August* | 78.8 | 77.6 | 71.3 | [50] |
| *13<sup>th</sup> August – 19<sup>th</sup> August* | 72.6 | 75.9 | 75.5 | [50] |
| *20<sup>th</sup> August-26<sup>th</sup> August* | 81.4 | 80.2 | 69.4 | [50] |

Our simulations over the period from September 2020 until December 2021 reflect a great deal of uncertainty, as can be seen from the shaded bands in Figures 3-4. Each of our model simulations represents one possible realisation of the flow of COVID-19 transmission among the population, but since this depends on the exact characteristics of who gets infected and when, the dynamics of transmission over long periods are subject to substantial uncertainties. While we have captured some of this uncertainty in our epidemic projections, we have not captured the impact of this on our estimates of the testing levels required to avoid a secondary epidemic wave, nor on our estimates of the population-level efficacy of masks. We have provided point estimates of the testing levels that would be necessary to avoid a secondary wave, but these should be interpreted with a degree of caution since they are based on the median simulation from a stochastic process with a degree of variation.

Finally, we note that we do not account for interactions between regions that differ in policy and infrastructure; for example the differences between Scotland’s ‘Test and Protect’ strategy and the NHS England ‘Test and Trace’ strategy. Rules on distancing and mask wearing have also been varied in different regions where prevalence is higher. We do not, however, attempt to model the effect of individuals moving between regions mediating the interaction of these different epidemics unfolding in different ways under different conditions. These aspects can be considered in future extension of this work.

Under the current policy on masks in UK, masks are not mandatory in all workplaces and hence we have not included these in our modelling framework. Furthermore, there are some variations in policy recommendations across the four UK nations with workers in some workplaces required to wear masks; for example workplaces that are open to the public requiring the workers to wear masks in Scotland but not in England [22]. Since our intention was to model the UK epidemic as a whole, we do not differentiate between these differences. Instead, we have assumed that up to 50% of the workforce will return to work, with the rest of the people remaining working from home, based on slight increases to the current estimates on the proportion of the workforce returning to work during July and August [49]. If policy changes and masks are made mandatory in workplaces, we could explore this scenario in future.

Similarly, we have modelled that in schools masks would be worn by secondary school children only since this is the policy recommendation informed by evidence that younger children have lower susceptibility and potentially lower transmissibility than adults [41]. In addition, we note that many of the school-based outbreaks observed globally have been concentrated in older students [44]. In the process of developing our models and preparing this manuscript, we have explored additional scenarios with masks mandatory in all schools, only primary schools etc, but the results of these other models are not included here as these are not under consideration for UK policy on masks. Within the modelling framework, we can evaluate a large number of scenarios considering different permutations of mask usage by different cohorts and across various settings. But the purpose of this study was to evaluate a feasible and policy-relevant subset of these permutations.

While our results report the COVID-19 cases/deaths projections over a relative long timescale, until the end of December 2021, we are aware that long-time forecasting with epidemiological modelling has a level of uncertainty. For example, future epidemic trajectories heavily depend on both human behaviour, future policy choices and future clinical therapies, which, as these are currently unknown, are not accounted for in the model. Specifically, this analysis does not assess future changes that may occur such as potential seasonality of the virus, reactive policies of, for example, intermittent local lockdowns or trade-offs in reopening society, or implementation of therapies to reduce severity of symptoms of COVID-19 related mortality or vaccination policies, thus making the model projections over the longtime period uncertain. But, since the scope of this analysis was to illustrate whether a resurgence of COVID-19 will occur later this or next year in a shape of a second epidemic wave, running the simulations until the end of 2021 was a necessity to illustrate this point and suggest when the potential peak may occur subject to the epidemic continuing on the current trajectory and under current conditions.

We also note that school-based reopening are expected to be accompanied by numerous countermeasures relating to physical distancing, hygiene, ventilation and ‘bubbles’ of year group in secondary schools and classes in primary schools as well as masks. With these countermeasures in place, it is expected that school-based transmission risk to students, teachers and staff may be somewhat mitigated, but the adherence to these policies by school children at all times is uncertain. Hence on balance, for this analysis, we have assumed that such measures could reduce transmission by 10%. We note that this is a modelling assumption and it may be an underestimation.

Furthermore, while our findings suggest that adequate TTI is required alongside masks wearing in secondary schools and in community settings to prevent COVID-19 resurgence, the increased numbers of contacts due to school mixing, and reduced adherence to school-based rules on social distancing, can easily overwhelm the TTI programs. The questions remain whether the UK has sufficient testing capacity to a) scale-up to levels suggested by our study and b) trace increasing numbers of contacts per index in schools, workplaces and community settings. We acknowledge that achieving this will be challenging, and especially so if the UK is faced with a second wave with possible test delays, test capacity being reached and the contact tracing system being overwhelmed. In addition, effective isolation is also important and the currently reported levels of isolation need to be improved [45-46]. All of these factors could further reduce the impact of the TTI programme.

### Implications for policy

Overall our findings suggest that mandating masks in parts of society and in secondary schools could be beneficial as it can reduce the strength of COVID-19 resurgence across the society. Wearing masks forms a barrier for the viral particles to pass from the wearer to people surrounding them and vice versa [42], and hence wearing them at schools could reduce COVID-19 incidence in students, staff and teachers. In future work, we plan to evaluate the trade-offs within UK society necessary to keep schools open as COVID-19 cases start to increase and project the contribution to the society COVID-19 transmission from different contact network layers and age groups.

### Conclusions

In summary, our modelling suggests that while adoption of masks in secondary schools in addition to community settings may contribute to reducing the size of a second wave, it is not sufficient to prevent a secondary COVID-19 wave in the UK. Instead, a masks policy would need to be combined with adequate TTI strategy that can test a large proportion of symptomatic people during their infectious period, effectively trace their contacts and isolate them.

## Methods

### Transmission model

Analogous to our recently published work [28], we modelled the spread of COVID-19 using Covasim, a stochastic individual-based model of SARS-CoV-2 transmission across a population. Development and implementation details can be found at http://docs.covasim.org with the methodology outlined in [36]. The model was previously applied to explore different scenarios of schools reopening in the UK [28], explore the epidemic spread in Australia [37] and explore different non-pharmaceutical interventions for epidemic control in Seattle, USA [36]. The code used to run all simulations contained in this paper is available from https://github.com/Jasminapg/Covid-19-Analysis.

For the purposes of the analyses presented in this paper, we used Covasim’s default parameters, pre-populated demographic data on population age structures and household sizes by country, and with four population contact network layers for schools, workplaces, households and community settings. The per-contact transmission probability (the risk of SARS-CoV-2 transmission during a contact between an infectious individual and a susceptible individual) is assumed to depend on the contact network. Covasim accounts for testing strategies via parameters that determine the probabilities with which people with different symptoms receive a test each day.

### Data sources and calibration

We used Covasim’s defaults to generate a population of 100,000 agents who interact over the four contact networks layers described above. To fit the model to the UK epidemic, we performed an automated search for the optimal values of the number of infected people on 21 January 2020, the per-contact transmission probability, and the daily testing probabilities for symptomatic individuals (P_s_) during May, June, July and August. The optimal values were the ones that minimised the sum of squared differences between the model’s estimates of confirmed cases and deaths, and data on these same two indicators between January 21, 2020 and August 28, 2020 collated from the UK government’s COVID-19 dashboard (https://coronavirus.data.gov.uk). These particular parameters were selected as the most important to estimate because of the considerable uncertainties around them. We accounted for effect of the lockdown by reducing the per-contact transmission probabilities from 23 March 2020, up to 2% of their pre-lockdown values within schools, and to 20% of their pre-lockdown values within workplace and community settings, and increased these in a phased way since the phased relaxing of the lockdown measures from 1^st^ June. Exact scaling is shown in Table 2.

**Table 2:**
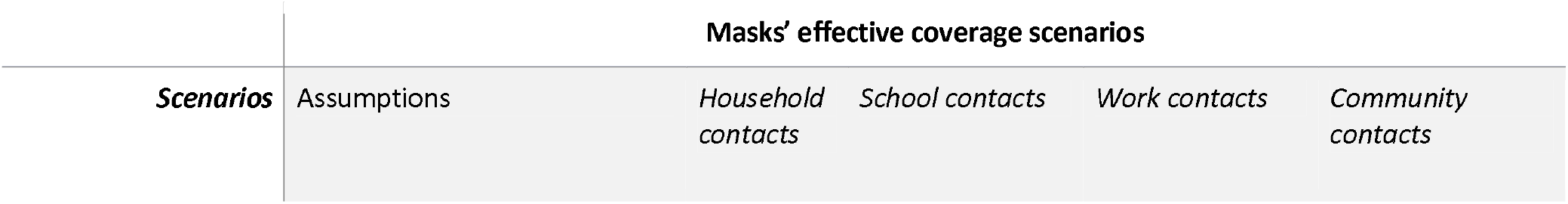
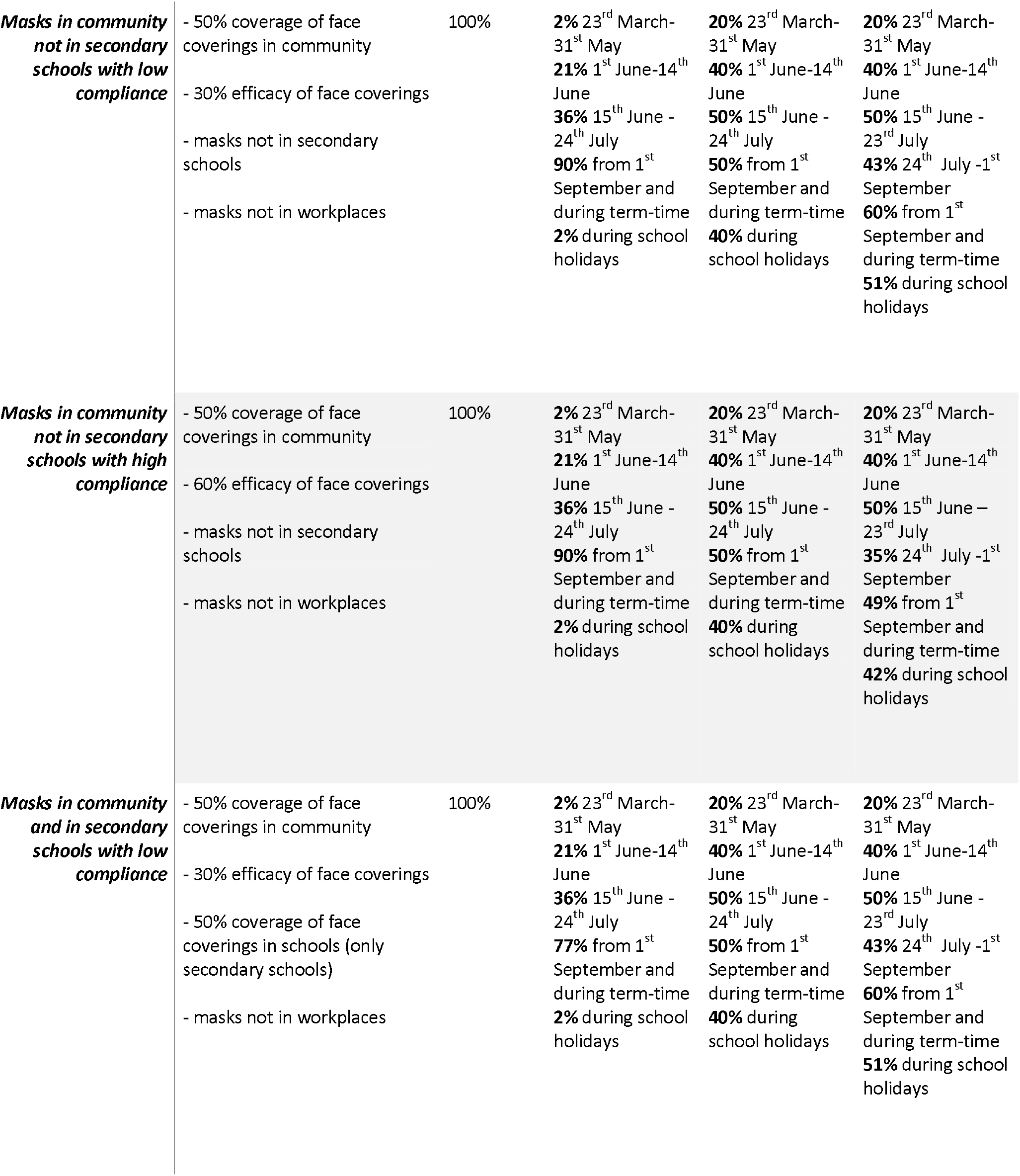
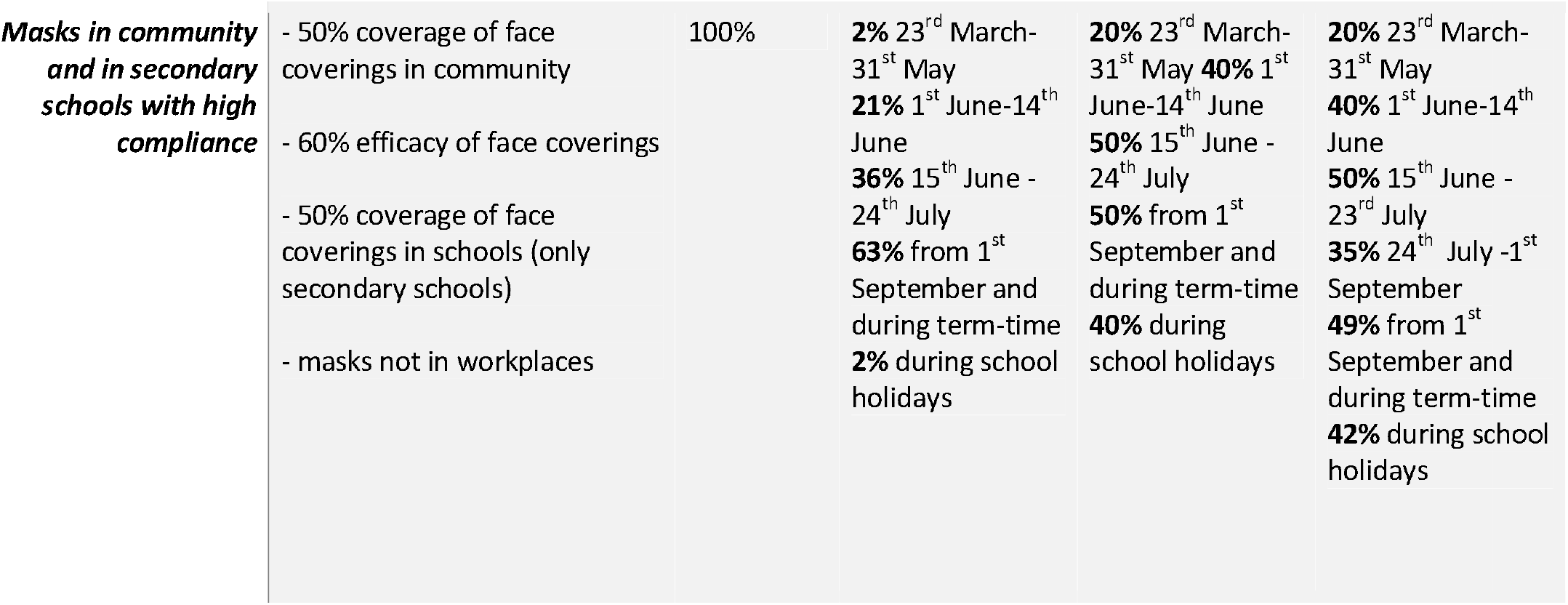
Scale factors applied to daily SARS-CoV-2 transmission probabilities in households, schools, workplaces, and the community under the scenarios of face coverings policy and compliance levels.

We also used publicly available weekly data from NHS Test and Trace to generate a level of contact tracing of contacts of those testing positive since the start of the programme on May 28, 2020 and collated in Table 1. We multiplied the percentage of people testing positive that were interviewed, the percentage of those reporting contacts and the percentage of contacts that were traced to generate an overall percentage for contacts of those tested positive that were traced. This assumed that all those testing positive would have the same number of contacts regardless of whether or not these were reported. We determined that this contact tracing level was 42% for June, 47% for July and 44% August, with an average of 47% tracing since 28^th^ May 2020. We assumed that 0.75% with asymptomatic COVID-19 infections being tested at some point during their illness (assuming an average symptomatic period of roughly 10 days) throughout June-August.

### Modelled effective coverage of different contact network layers

In this study we used effective coverage as a measure for effectiveness across all types of face coverings, and we translated this into the modelling framework by reducing the transmission probability of different population layers.

For the policy, we assumed that either masks will be mandatory in parts of community, such as public transport from June 15, 2020, and then extended to more places, such as shops, from July 24, 2020, but not in schools from 1^st^ September, or that masks will be mandatory in parts of community from June 15, 2020, and then in more places from July 24 and also in the communal areas of secondary schools outside of the classroom from 1^st^ September. For each scenario, we derived effective coverage of face coverings as a product of efficacy and coverage (proportion of contacts in which at least one mask is used) across layers. We modelled these policies by scaling down the probability of transmission by this effective coverage level in the relevant layers depending on the two policies of face coverings with details in Table 2. We used two values of effective coverage to describe a lower and higher effective coverage of face coverings across settings and below we describe how we arrived at these two values.

#### Efficacy of masks

The efficacy of masks was defined as the size of the reduction in transmission probability during a contact between a susceptible and exposed individual when a mask is worn by one or both parties. For protecting the healthy wearer, systematic reviews and meta-analyses in other viruses have found mean effect sizes of around 45% in community settings [12] with a range of approximately 20–80% [12-14]. These should be adjusted downwards to account for different mask types, since cloth masks, the kind most commonly used among the UK public [7] are less effective than the N95 ventilators used by some study participants [12]. The reviewed studies, mostly case-control, were also subject to potential biases that may have inflated the effect size; for example, people who wear masks may tend to be more careful, for example in terms of washing hands more often and in keeping greater distance from cases, than those who do not. However, we must also factor in source control. Experiments have shown face coverings, especially but not exclusively medical masks, to greatly reduce viral shedding and droplet dispersal [8-9]. Taking all of these in consideration, we modelled a mean of 45% with a range of 25–70% as a reasonable estimate of face covering efficacy. This is calculated as the weighted average of one-person masking and both-people masking.

#### Coverage of masks across different contact network layers

##### School contact network layer

Within the model, we had to make assumptions on who may be wearing masks at schools. The education system in the UK consists of 14 school years (age 4-18 years) each of which start in September and finish the following July. There are seven years of primary school, with children entering reception aged 4 and leaving aged 11, followed by seven years of secondary school. Following discussion among the authors and with scientific advisors and policy decision makers in the UK, we decided to model the use of masks by secondary school students only, i.e. the last seven years of education. This implies that the coverage of masks in the school layer is at most 50%. The current masks policy is that masks are recommended in corridors and other communal “hot spots’ areas where there is a higher risk of COVID-19 transmission [47]. But this is implemented differently across different schools, and for simplicity we have assumed that the coverage of face coverings in the school layer is at most 50%. We note that this would be reduced if that that masks are not worn in classrooms and if students’ use of masks is not perfect. As before, the efficacy of face coverings was estimated to be 25–70%, and to account for uncertainty, we modelled two efficacy values of 30% and 60%. Multiplying by a coverage level of 50% we derived an effective coverage of 15% and 30% describing lower and higher effective coverage of masks in schools.

##### Community contact network layer

In our previous work [28] we assumed that with schools reopening, the society will reopen proportionally to school years going back. This would imply that with schools fully going back, the society would reopen to 90% of its pre-COVID-19 level. However, recent reports from surveys tracking the increase in contact rates suggest that during July and August the contact rate has only marginally increased to at most 4 contacts per person which is 36% of the pre-COVID-19 level. This is anticipated to increase with schools reopening but we anticipate not to 90% as modelled before, but likely to around 70% of the pre-COVID-19 level.

In terms of mask usage, in the UK, self-reported surveys for August 31-Sep 06 suggests that 64% of Britons “always” and 15% of Britons “frequently wear mask while in public places [48]. Since we do not model specific parts of the community, e.g. transport network or shops, separately, we use a slightly larger than the mean coverage level reported in the surveys [48] and assume that overall adherence to masks in community is around 50%. As before, the efficacy of face coverings was estimated to be 25–70%, and to account for uncertainty, we modelled two efficacy values of 30% and 60% producing effective coverage levels of 15% and 30%.

##### Workplace contact network layer

Face coverings are not currently mandatory in workplaces across UK. In our previous work [28], we assumed that with schools reopening, 70% of workplaces will also reopen to al staff. A recent report from the Office for National Statistics suggests that 49% of workers are working from home [49]. We anticipate the proportion of people going back to work to increase when schools reopen. We thus assume across scenarios that 60% of the workforce will return with reopening of schools during school term-time, 10% of which will additionally work from home during the school holidays. We also assume that making the workplaces COVID-19 secure will also reduce transmission in the workplace, and hence model the transmission probability for the workplace layer to be 50% of the pre-COVID-19 level during term time and 40% during school holidays.

#### Scenarios

While it is possible in our framework to explore the effect of varying assumptions regarding mask type, effectiveness, compliance, geographical location, setting (school, workplace, community) and other factors that may affect the impact of mask policies, this would require sampling an infeasible number of scenarios, and be of limited value given the standard of evidence available. Instead, under the scenarios of reopening of schools from September 2020, alongside society, and under different levels of TTI strategies, we simulated four scenarios describing two policies on face coverings and two levels of face coverings effective coverage. The four scenarios are modelled by reduction in the transmission probability across layers as described in Table 2.

#### Analysis

For each of the four scenarios, we estimated the daily and cumulative numbers of infections and deaths, until 31 December 2021 for all possible combinations of test and trace levels. Since Covasim is stochastic, we simulated each scenario under 12 different random number seeds, and we present the median estimates along with ranges corresponding to the upper and lower bounds generated by these 12 seeds.

Across each scenario, we illustrated our results for two test-trace combinations using the current tracing level and two testing levels: current and scaled-up to avoid a second wave.

## Data Availability

The data that support the findings of this study are available from the corresponding author upon reasonable request.

## Code availability

The code used to run all simulations contained in this paper is available from https://github.com/Jasminapg/Covid-19-Analysis.

## Acknowledgements

We would like to acknowledge Prof Graham Medley (LSHTM) and Dr Edwin van Leeuwen (Public Health England) for helpful discussions around the modelling scenarios. WW acknowledges the support of the Chief Scientist Office (COV/EDI/20/12).

## Author contributions

JPG conceived the study and developed the specific modelling framework, based on the Covasim model developed by CCK, RMS, DM and DJK. JPG, DF, WW, CCK, RMS, DM and DJK collated data for the parameters used. JPG ran the modelling analysis with input from WW, CCK, DM and RMS. JPG, CB and RV defined the different scenarios in the UK context following conversations with scientific advisors within SPI-M and SPI-B. JPG wrote the manuscript with input from CB, RV, WW, CCK, RMS, DM, DF and DJK. All authors approved the final version.

## Material & Correspondence

Correspondence to Jasmina Panovska-Griffiths

## Supplementary material

**Figure S1:**
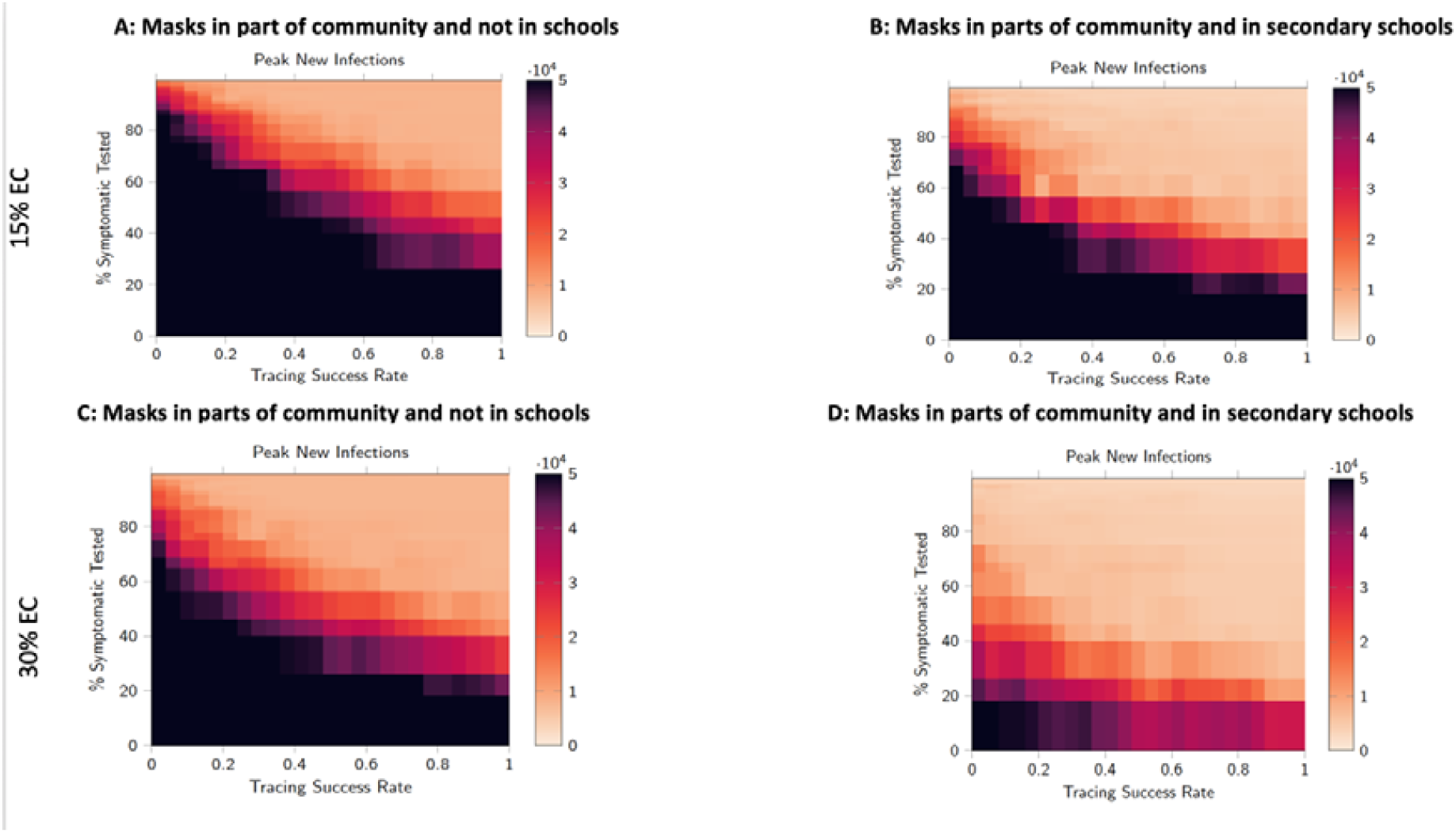
Heatmaps of the peaks of new infections for different trace (x-axis) and test (y-axis) levels across the scenario of masks wear in parts of community with and without schools masks’ wear. Higher values of the peak of new infections are shown in darker shades of red, while lower values are lighter colours with a light orange colour representing a region within where the resurgence of COVID-19 is controlled with combinations of adequate test-trace and mask wear.

**Figure S2:**
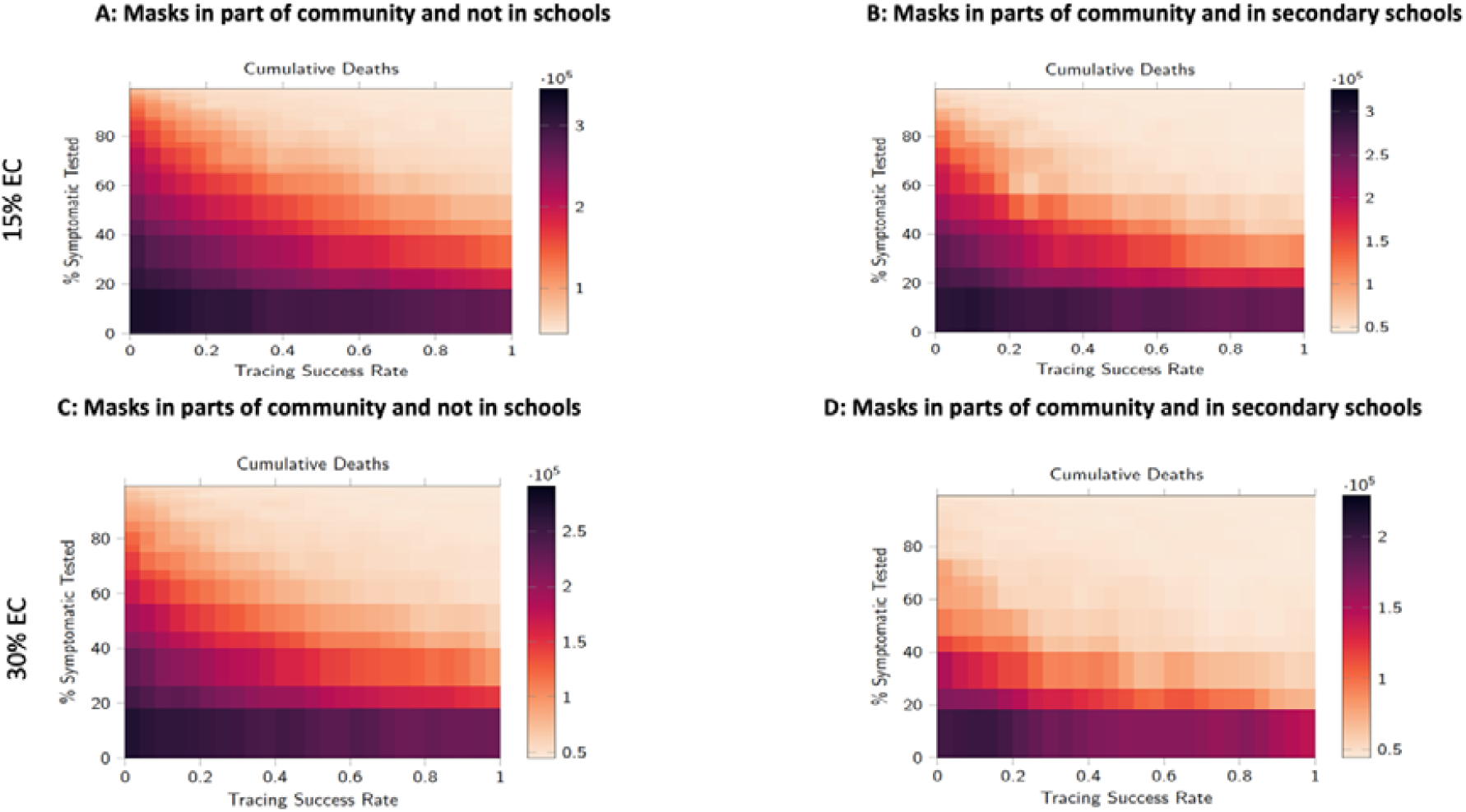
Heatmaps of the cumulative deaths for different trace (x-axis) and test (y-axis) levels across the scenario of masks wear in parts of community with and without schools masks’ wear. Higher cumulative deaths are shown in darker shades of red, while lower values are lighter colours with a light orange colour representing a region within where the resurgence of COVID-19 is controlled with combinations of adequate test-trace and mask wear.

**Table S1:** Parameters fitted during the calibration.

| Calibration of the model |  |
| --- | --- |
| Initial seeding of infectious persons on 21/02/2020 | 1500 |
|  | 0.0071967 |
| in March | 0.009 |
| in April | 0.013 |
| in May | 0.026 |
| in June | 0.0277 |
| in July and August | 0.0277 |
| in May and June | 0.00075 |

**Table S2:** Disease duration parameters, in days, used in simulations. This table is modified from [36], which describes the Covasim model in more detail.

| Disease Duration Parameters |  |  |  |
| --- | --- | --- | --- |
| Parameter | Description | Distribution (mean, std) | Source |
|  | Length of time after exposure before an individual is infectious (i.e. has begun viral shedding) |  | [1-5] |
| $i_1$ | Length of time after viral shedding has begun before an individual has symptoms | $i_1 \sim \text{lognormal} (1, 0.9)$ | [1,7] |
| $i_2$ | Length of time after symptoms appear before they become severe and the person requires critical care | $i_2 \sim \text{lognormal} (6.6, 4.9)$ | [7] |
| $i_3$ | Length of time after severe symptoms appear before the person requires critical care | $i_3 \sim \text{lognormal} (3, 7.4)$ | [8] |
| $r_a$ | Recovery time for asymptomatic cases | $r_a \sim \text{lognormal} (8, 2)$ | [6] |
| $r_m$ | Recovery time for mild cases | $r_m \sim \text{lognormal} (8, 2)$ | [6] |
| $r_s$ | Recovery time for severe cases | $r_s \sim \text{lognormal} (14, 2.4)$ | [5] |
| $r_c$ | Recovery time for critical cases | $r_c \sim \text{lognormal} (14, 2.4)$ | [5] |

**Table S3:** Age-linked disease parameters used in simulations. This table is borrowed from [36] describes the Covasim model in more detail. Key: *p*_*sus*_: odds ratio of developing symptoms; *p*_*sym*_: probability of developing symptoms; *p*_*sev*_: probability of developing severe symptoms (i.e., sufficient to justify hospitalization); *p*_*cri*_: probability of developing into a critical case (i.e., sufficient to require ICU); *p*_*death*_: probability of death.

| Parameter | Age-linked Disease Parameters |  |  |  |  |  |  |  |  |
| --- | --- | --- | --- | --- | --- | --- | --- | --- | --- |
|  | 0-9 | 10-19 | 20-29 | 30-39 | 40-49 | 50-59 | 60-69 | 70-79 | 80+ |
| $p_{sus}$ | 0.34 | 0.67 | 1.00 | 1.00 | 1.00 | 1.00 | 1.00 | 1.24 | 1.47 |
| $p_{sym}$ | 0.50 | 0.55 | 0.60 | 0.65 | 0.70 | 0.75 | 0.80 | 0.85 | 0.90 |
| $p_{sev}$ | 0.0005 | 0.0065 | 0.0072 | 0.0208 | 0.03430 | 0.0765 | 0.1328 | 0.2066 | 0.2457 |
| $p_{cri}$ | 0.00003 | 0.00008 | 0.00036 | 0.00104 | 0.00216 | 0.00933 | 0.03639 | 0.08923 | 0.1742 |
| $p_{death}$ | 0.00002 | 0.00006 | 0.00030 | 0.00080 | 0.00150 | 0.00600 | 0.02200 | 0.05100 | 0.09300 |
| $\beta$ | 1.00 | 1.00 | 1.00 | 1.00 | 1.00 | 1.00 | 1.00 | 1.00 | 1.00 |

